# The impact of COVID-19 upon the delivery of exercise services within cystic fibrosis clinics in the United Kingdom

**DOI:** 10.1101/2021.04.12.21255205

**Authors:** Owen W Tomlinson, Zoe L Saynor, Daniel Stevens, Don Urquhart, Craig A Williams

## Abstract

The COVID-19 pandemic has resulted in unprecedent change to clinical practice. As the impact upon delivery of exercise services for people with cystic fibrosis (CF) in the UK was unknown, this was characterised via a national survey. In total, 31 CF centres participated. Principal findings included a significant reduction in exercise testing, and widespread adaptation to deliver exercise training using telehealth methods. Promisingly, 71% stated that they would continue to use virtual methods of engaging patients in future practice. This does, however, highlight a need to develop sustainable and more standardised telehealth services further to manage patients moving forwards.

## 1. INTRODUCTION

Participation in regular physical activity (PA) and exercise is of benefit for people with cystic fibrosis (CF), and is an integral part of their clinical management. Regular exercise testing and reviews of exercise training programme are therefore recommended to occur on at least an annual basis [1, 2].

Upon the emergence of the SARS-CoV-2 coronavirus-2019 (COVID-19), restrictions were imposed by the UK Government to limit transmission, and included additional constraints for clinically vulnerable individuals, such as people with CF [3]. Specifically, people with CF were asked to ‘shield’ at home, with recommendations to also avoid face-to-face hospital appointments where possible [4]. Early evidence from Switzerland reported that such measures had a negative impact upon the PA levels of people with CF, with noted barriers including closure of facilities, lack of motivation, and cancelled training supervision [5].

As with many services within the National Health Service (NHS), CF multi-disciplinary care teams (MDTs) have been forced to adapt and deliver services virtually where possible (e.g. telephone, video and email consultations) [6]. However, it is unknown how feasible such changes are for CF MDTs, and to what extent exercise services in particular have been affected. Given the importance of exercise testing and training for people with CF, identifying how exercise services have changed during the COVID-19 pandemic is a priority; to ensure high quality services are still delivered, and identify areas where additional resources are required.

Understanding service change does however have applicability beyond the current pandemic. Given that people with CF are regularly advised to segregate from one another for infection control reasons, and ‘shield’ at home when unwell, any positive or sustainable changes to clinical practice may be appropriate for continued implementation. This online survey therefore sought to identify the impact of the COVID-19 pandemic upon the delivery of exercise services within CF clinics in the UK.

## 2. METHODS

Questions asked within this survey formed part of a wider survey related to exercise services in CF MDTs across the UK, in a replication of previous work [7]. Information concerning centre location, patient population, and job role of respondents was collated, alongside specific COVID-19 related questions provided in Supplementary File 1.

The survey was distributed via email, by the Association of Chartered Physiotherapists in CF, the UK CF and Exercise Technicians Network, and UK CF Medical Association to their respective memberships. It was asked that a single member of each team (ideally the person responsible for exercise services, regardless of position) completed the survey on behalf of their site, to ensure a single response per centre. The survey link was distributed in January 2021 and remained open for 6 weeks, to maximise the response rate. This survey was hosted on an online platform (Qualtrics XM; Provo, Utah, USA), chosen because of its compatibility with both computers and smartphones, whilst also ‘whitelisting’ IP addresses for compliance with data protection regulations.

This study was approved by the University of Exeter Sport and Health Sciences Ethics Committee (200708-A-01). All respondents provided consent to participate via a series of check-boxes, confirming they had read and understood the participant information sheet and were providing information on behalf of their centre.

Data is presented as frequency statistics, and free-text responses are provided to emphasise predominant themes within responses.

## 3. RESULTS

The survey was completed by *n* = 31 respondents from across the UK. This represented specialist (*n* = 24; ∼50% of specialist UK centres) and network (*n* = 7) centres, covering adult (*n* = 11), paediatric (*n* = 16) and mixed (*n* = 4) care centres. In total, *n* = 27 respondents were Physiotherapists (lead CF specialist, *n* = 15; CF specialist, *n* = 11; non-CF specialist, *n* = 1) within their respective CF MDTs. The remaining respondents were completed by Therapy Assistant/Technicians (*n* = 2), Exercise Therapist (*n* = 1) and Exercise Practitioner (*n* = 1).

The majority of respondents stated the pandemic had restricted their ability to undertake exercise testing (97%) and exercise training (71%), with the relative frequency of both being negatively affected, as shown in Figure 1.

**Figure 1.**
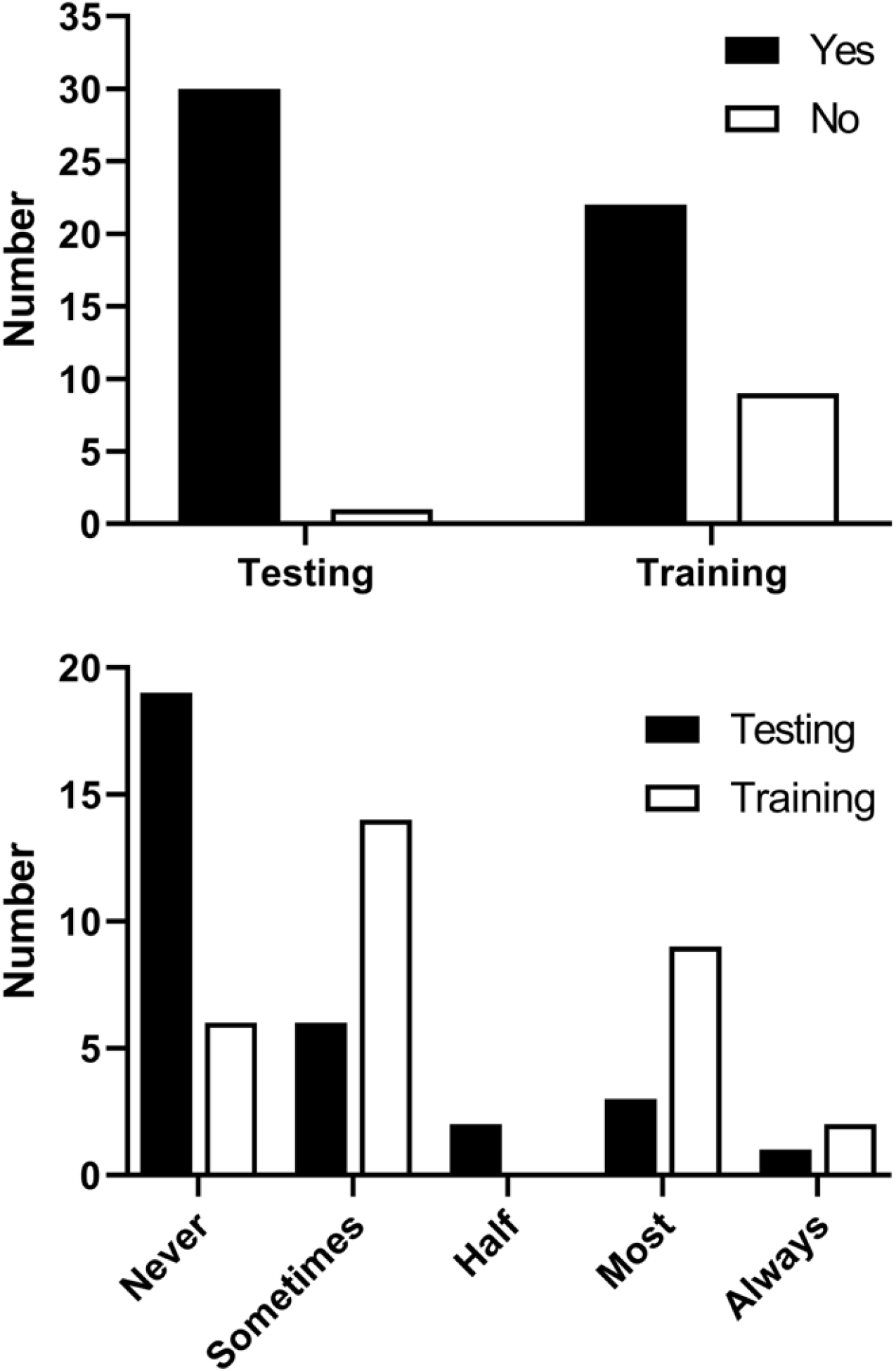
Number of responses to questions 1 and 4 (*Has the COVID-19 pandemic affected your ability to deliver exercise testing/training?*) and 2 and 5 (*How often are you able to undertake exercise testing/training due to the pandemic?*).

Free-text responses to questions related to adaptation of practices, barriers and beneficial resources highlighted a number of common themes, as shown in Table 1. A large proportion of respondents (*n* = 22, 71%) stated that they would maintain some form of telehealth (e.g. delivery of classes, home monitoring and consultations) within their clinical practice (free-text responses are provided in Table 2).

**Table 1.** Common themes highlighted in free-text responses to questions related to adaptation, barriers, and beneficial resources.

| Question | Responses |
| --- | --- |
| How has your centre adapted exercise testing in light of the pandemic? | Stopped altogether ($n = 23$)<br>Inpatient testing only ($n = 6$)<br>Virtual/home-based testing ($n = 5$) |
| How has your centre adapted exercise training in light of the pandemic? | Video/online classes/sessions ($n = 22$)<br>Inpatient training ($n = 5$)<br>Stopped altogether ($n = 2$)<br>Home-visits ($n = 1$)<br>Outdoor exercise ($n = 1$) |
| What have been the major barriers to delivering exercise services (testing and training) during the pandemic? | Access to patients ($n = 12$)<br>Reduced staffing ($n = 10$)<br>Access to facilities/space ($n = 9$)<br>Concern around exercise as an aerosol generating procedure ($n = 5$)<br>Local and national restrictions (e.g. movement/home visits/distancing) ($n = 5$). |
| What resources have you found to benefit your team in during the pandemic? | Improved IT facilities and access to online platforms ($n = 13$)<br>Existing online classes/resources ($n = 10$)<br>Additional finances made available for equipment ($n = 3$) |

A full list of free-text responses is provided in Supplementary File 2.

**Table 2.** Selected free-text responses to question 9: “Are there any changes you have made due to the pandemic that you intend to keep and/or maintain?”, with telehealth highlighted as a predominant thematic response. Responses are provided as written by respondents, with only spelling mistakes changed to increase readability. A number of single word responses (e.g. *“videos”*) have been omitted, but counted in statistics. Free-text responses with identifying information have been removed.

|  |  |
| --- | --- |
| **Telehealth** | *“Continue doing exercise classes through video.”*<br>*“IT and video consultation support.”*<br>*“Continue to do online live and on demand classes for the foreseeable future.”*<br>*“More virtual exercise sessions with patients at home via Attend Anywhere.”*<br>*“Virtual Leisure Centre classes.”*<br>*“Some video calls for patients who can't attend in person.”*<br>*“Virtual exercise support.”*<br>*“We will keep doing virtual classes.”*<br>*“Video consultations, remote lung function monitoring.”*<br>*“More virtual/video reviews as needed.”*<br>*“Video clinics and spirometry.”*<br>*“Yes, the video sessions have been a great success.”*<br>*“Will keep virtual exercise training sessions.”*<br>*“Video monitoring and calling.”*<br>*“Some video conferencing. This enables you to see the house and be realistic about what can be done in the home.”*<br>*“Retain video calls.”*<br>*“Video calls for all patients.”*<br>*“Using video calling systems more in the future.”*<br>*“Keep: - video clinics and exercise reviews - Chester Step Test as an extra exercise testing modality.”*<br>*“Continue with virtual sessions. Hoping to provide group sessions.”*<br>*“Would continue with some patients doing exercise sessions via video calls.”* |
| **Other Responses** | *“Continue with most of changes, hope to reintroduce exercise testing. Looking at sit to stand test.”*<br>*“I would intend to keep the weekly exercise programming going maybe in school holidays or especially while gyms are shut, but redeployment will not allow this to continue, but could be reinstated when we start to return to normal.”*<br>*“Polar coach. Looking to set up team Strava account if possible in line with trust regulations. We are also looking into the use of social media to help promote exercise and our exercise groups.”*<br>*“None from an exercise viewpoint.”* |

Patients asked their MDTs a variety of questions about exercise and CF, with safety surrounding exercise and ideas for exercises cited as common questions (Supplementary File 2). The majority of respondents (*n* = 27) felt they were able to confidently answer these questions. Respondents themselves highlighted a number of questions and comments, across numerous themes, including short-term and long-term care for people with CF, guidelines for ongoing practice, and the changing nature and direction of engagement with exercise (Supplementary File 2).

## 4. DISCUSSION

This is the first evaluation of how the COVID-19 pandemic has impacted the delivery of exercise services in UK-based CF centres. Our findings demonstrate that, whilst exercise testing services have been drastically reduced, exercise training provision has adapted, and has continued to be offered in novel ways, particularly through an increased use of digital health technology.

It is clear from our representative group of CF centres that many have adapted their clinical practice in multiple ways, with routine use of video-calling noted as a predominant theme. The use of telehealth to deliver exercise training for people with CF prior to the pandemic was previously shown to be feasible [8], and delivery of exercise services using virtual platforms during the pandemic has since been described anecdotally [9] and evaluated [10] within single CF centres in the UK. The present survey shows promising implementation of this practice for the benefit of people with CF, and further notes that many centres plan to continue to utilise online resources for clinical practice.

Aside from exercise services, telehealth has emerged as a promising tool for delivery of CF care, with single-centre descriptions of changing practice detailing acceptance and challenges of such an approach [11, 12]. This adoption of digital services aligns with the long-term NHS strategy of utilising more technology in routine care [13], as well as the recent ‘Carter Report’; a review of efficiency and productivity within the NHS, that highlighted the need for enhanced digital solutions [14]. The present findings that CF MDTs are currently adapting practice, and using digital tools with a view to long-term adoption of these services is therefore encouraging.

However, as noted within free-text responses to this survey, CF MDTs face continued challenges concerning finances, staffing, equipment, and space in order to ensure high-quality services are maintained. Therefore, individual CF MDTs, NHS Trusts and Clinical Commissioning Groups must be aware of, and adequately address, these anticipated challenges to ensure successful continuation of adapted services.

## Data Availability

Data not available due to ethical restrictions.

## ABBREVIATIONS

CF: cystic fibrosis
MDT: multi-disciplinary team
NHS: National Health Service
PA: physical activity
UK: United Kingdom

## ACKNOWLEDGMENTS

The authors would like to thank Dr Caroline Elston, Thomas Kent, Lisa Morrison, and Dr James Shelley for assistance with distribution of the survey, and all respondents for their time in completing the survey.

## FUNDING

This survey was undertaken as part of the ‘Youth Active Unlimited’ Strategic Research Centre (SRC), funded by the Cystic Fibrosis Trust (SRC#008). CA Williams is the Chief Investigator of this SRC.

## CONFLICTS OF INTEREST

There are no conflicts of interest to report.

## CONTRIBUTION STATEMENT

All authors conceived and designed the study; OWT and CAW coordinated delivery of survey and collation of results; OWT analysed results and drafted the manuscript; all authors critically revised and approved final manuscript for publication.

## Notes

### Competing Interest Statement

The authors have declared no competing interest.

### Author Declarations

This study was approved by the University of Exeter Sport and Health Sciences Ethics Committee (200708-A-01).

