## Supplementary File 1 for "The impact of COVID-19 upon the delivery of exercise services within cystic fibrosis clinics in the United Kingdom"

**Supplementary File 1.** Survey questions related to COVID-19 and exercise services

| | |
|---|---|
| **Q1** | Has the COVID-19 pandemic affected your ability to deliver exercise testing?<br>YES NO |
| **Q2** | How often are you able to undertake exercise testing due to the pandemic?<br>ALWAYS MOST OF THE TIME ABOUT HALF THE TIME SOMETIMES NEVER |
| **Q3** | How has you centre adapted exercise testing in light of the pandemic (e.g. video tests, home visits, stopped altogether, no change)* |
| **Q4** | Has the COVID-19 pandemic affected your ability to deliver exercise training?<br>YES NO |
| **Q5** | How often are you able to undertake exercise training due to the pandemic?<br>ALWAYS MOST OF THE TIME ABOUT HALF THE TIME SOMETIMES NEVER |
| **Q6** | How has you centre adapted exercise training in light of the pandemic (e.g. video tests, home visits, stopped altogether, no change)* |
| **Q7** | What have been the major barriers to delivering exercise services (testing and training) during the pandemic?* |
| **Q8** | What resources have you found to benefit your team in during the pandemic?* |
| **Q9** | Are there any changes you have made due to the pandemic that you intend to keep and/or maintain?* |
| **Q10** | What questions have your patients been asking you in relation to exercise and COVID-19?* |
| **Q11** | Have you been able to confidently answer your patients questions?<br>YES NO NO PATIENTS HAVE ASKED QUESTIONS |
| **Q12** | Do you have any questions with regards to exercise and COVID-19 for cystic fibrosis that you would like answering/addressing?* |
| **Q13** | Do you have any final comments on exercise and COVID-19 in your centre?* |

*Indicates questions were free-text responses.
