## Supplementary File 2 for "The impact of COVID-19 upon the delivery of exercise services within cystic fibrosis clinics in the United Kingdom"

**Supplementary File 2.** Free-text responses to questions 3, 6, 7, 8, 10, 12, and 13.

| **How has you centre adapted exercise testing in light of the pandemic?** |
|---|
| *"Stopped annual exercise testing; but no clinical risk to cohort, therefore not sourced alternative ways to test."* |
| *"Unable to do modified shuttle walk test, have started doing Chester step test. Only do testing face to face if video call unable to do test."* |
| *"Forced stop due to staff limitations, restrictions in hospital attendance and for AGP with outpatients."* |
| *"Not able to do formal testing with most patients, but if they are admitted for any reason we can take the opportunity to do a field test."* |
| *"Continued with inpatient testing, virtual testing via video link. Overall large decrease in testing due to the pandemic."* |
| *"Only on inpatients if well enough."* |
| *"We have not completed any of our usual incremental step tests since the start of COVID-19. We are aiming to re-start them at annual assessments and as outpatients from January 2021."* |
| *"Unable to deliver due to being moved out of hospital grounds – no space available."* |
| *"Able to carry out 6 minute exercise test, but only on inpatients and these have to be outdoors. CPET completely stopped."* |
| *"Dependent on government guidelines at any time, some tests carried out at home. Occasionally managing to test at clinic appointment."* |
| *"CPET stopped altogether. Completing sit to stand virtually or face to face if required."* |
| *"Stopped all together routinely. Only step tests completed if patient is an inpatient as education/assessment tool or to show progression during course of IV."* |
| *"Moved it outside to a covered carpark, so can only be done when not too cold and not at busy car park times."* |
| *"No CPET or Shuttle tests during COVID. Tests at home face to face 3 minute step, IST, Functional Strength tests. Video reviews and tests as well."* |
| *"Stopped altogether as all reviews are virtual. We had only just got our new bikes set up so are using the time to ensure we get the best set up of those. We were ready to roll this out just prior to lockdown 3.0."* |
| *"Considering the STS60 however with most staff redeployed this is very difficult."* |
| *"Testing was stopped for some time. Just getting some fitness testing (CPET) back, although field testing has not started again yet."* |
| *"Inpatient numbers down from 13-15 to max of 4 due to bed availability. Inpatient exercise test - Chester Step Test as interim monitoring exercise testing tool with good effect. No outpatient exercise testing currently."* |
| *"Stopped altogether currently but hoping to start STS virtually and back to community as able."* |
| *"At present stopped altogether as most contacts between physio staff and patients are either telephone or video calls. No out patient service available and no MDT clinics running face to face."* |

| How has you centre adapted exercise training in light of the pandemic? |
|---|
| *"No direct impact from Pandemic as wasn't completing prior to."* |
| *"Started doing exercise classes via video."* |
| *"More shift to video calls and online exercise programmes."* |
| *"Advice and suggestions online."* |
| *"Online Live exercises 4 x weekly. This has improved as a consequence of the Pandemic as prior to this we did not offer this option."* |
| *"We are still doing inpatient exercise sessions. We continue to assess and discuss exercise in clinics and at annual review and offer advice via virtual clinics on how to increase exercise and encourage."* |
| *"Virtual appointments, virtual group sessions, email communication with patients regarding exercise, clinic drop ins virtually."* |
| *"Only able to see inpatients for this, and then can only take inpatients out for a walk around the hospital site, or use a static bike with them within their inpatient cubicle."* |
| *"We have provided information on exercise available online - Joe Wicks, Oti Mabuse dance classes, Cosmic Kids Yoga etc. Inpatients were limited to exercising in their rooms, but can now use our gym once a negative COVID test is confirmed. We have offered video reviews for exercise support as needed."* |
| *"Home visits and outdoor exercise, but this is limited."* |
| *"Limited by space restrictions. Have increased video exercise sessions."* |
| *"We ran a first lockdown exercise challenge on a weekly basis for 5 weeks, patients were all given an exercise programme based on their age and ability to carry out at home as many times as they liked in the week - each session was around 10-15 mins."* |
| *"Delivering virtual (video) exercise sessions and supporting patients via telehealth.2* |
| *"Giving out exercise programmes, videos on YouTube, setting stair challenges in the home during shielding etc. Monitoring with video calls and at clinics."* |
| *"Encouragement via video link and ideas of programmes to follow that are web based. Can still use the small on site gym for one patient per day."* |
| *"Yes, more exercise testing after first Lockdown and introduction of Genetic Modifiers"* |
| *"We offer 2 x week virtual sessions still, and have supported many of our patient with a weeks' worth of physio support doing BD sessions often incorporating exercise training into."* |
| *"Discussion via video clinics and emailing programmes."* |
| *"Video calls, online classes, outpatient exercise prescription."* |
| *"Video clinics proved useful, still delivering exercise training for outpatients through this mainly pre-transplant. Inpatient - gym area not available since Pandemic. Providing equipment for patients rooms. Not allowed to leave hospital premises to exercise outside."* |
| *"Video calls for exercise training. As we know patients well this has been relatively easy to do as have knowledge of patients exercise ability. Have carried out some group sessions with maximum of 4 patients joining video call for exercise. Try to ensure patients have similar exercise ability."* |

| What have been the major barriers to delivering exercise services (testing and training) during the pandemic? |
|---|
| *"Space access, adhering to COVID recommendations."* |
| *"Foot fall limitations on site. Concerns around AGP. Limited staffing and access to equipment."* |
| *"Staff redeployed, no rooms, patients unwilling to attend, equipment."* |
| *"Access to patients."* |
| *"Not seeing patients FTF initially."* |
| *"For testing - space. Cannot be done as safely and as accurately virtually. Change type of test selected e.g. 6MWT to 1 min STS test to be able to do this virtually/over video link with patients."* |
| *"Losing gym space for some time. Not being allowed."* |
| *"Frequent changes in NHS guidelines."* |
| *"IT security limiting virtual classes initially."* |
| *"Considered an AGP, patients shielding so needing to remain isolated, time (reduced staffing due to shielding, isolation, redeployment).!* |
| *"Closure of our paediatric gym preventing inpatients using it. Inpatients not allowed to leave their side-room, so limited space to exercise during admission. More virtual clinics therefore less opportunities for exercise testing. Exercise testing resulting in an AGP (coughing) and would therefore prevent further use of the room in clinic for a few hours. Difficult to deliver in the community due to lockdown restrictions and initial shielding."* |
| *"Time/staff/patient engagement."* |
| *"Unable to test, reduced space, staff being redeployed, unable to carry out home visits."* |
| *"Social distancing restrictions. Face to face contact kept to minimum."* |
| *"Space, time and staff, and justification for bringing people up to the hospital for a non-essential procedure."* |
| *"Had to move out of main CF centre due to COVID. Unable to access preferred method of assessment. Limiting face to face assessments with patients."* |
| *"Staff redeployed, unable to get patients in."* |
| *"Patient preference to avoid hospitals unless absolutely necessary, and cessation of outpatient services."* |
| *"Staffing, cross covering other areas, all non-urgent appointments were cancelled."* |
| *"Infection control procedures and clearing rooms after use."* |
| *"Same as pre-COVID."* |
| *"Local COVID-19 policy."* |
| *"No CPET or Shuttle tests due to decreasing footfall in hospital."* |
| *"Exercise and airway clearance is an AGP, so would require full PPE - not always appropriate."* |
| *"Not seeing patients face to face."* |
| *"Unable to bring patients to hospital, redeployed staff."* |

*"Patients unable to attend the hospital."*

*"Limited patient numbers for inpatients, not being able to see patients at home/in community. Usual area for exercise tests not available."*

*"Time allowed for virtual delivery."*

*"Staff redeployment. No face to face contacts. Some patients do not want video sessions."*

**What resources have you found to benefit your team in during the pandemic?**

*"Video call websites, WhatsApp, Microsoft teams."*

*"Enhanced IT support with a move to video consultation."*

*"YouTube videos, beam, CFyogi, Joe Wicks."*

*"Online opportunities, opportunity to access Beam feel good and use their platform to deliver our own Live classes."*

*"Beam, Zoom, patients having home equipment e.g. exercise bikes, therabands, email/IT."*

*"Online training for virtual training."*

*"Online exercise for children - Joe Wicks PE, Cosmic Kids Yoga, Oti Mabuse Dance Classes. Hospital charity funding to get more equipment for use in the side rooms on our ward. More laptops to enable delivery of video reviews."*

*"Video links. Home spirometry and monitoring."*

*"Use of 'attend anywhere' for virtual sessions. Recommending multiple pre-recorded exercise sessions for patients to use at home."*

*"Joe Wicks PE. Any online exercise facilities."*

*"YouTube videos, video calling to monitor."*

*"Charity funding of home exercise equipment. Less staff in car park so able to use as an outside exercise area."*

*"CF Trust equipment grants."*

*"Online exercise resources (Joe Wicks, BEAM, kids exercise Ninja videos on YouTube)."*

*"Improved familiarity with MS Teams for virtual exercise classes and virtual support weeks. Improved functionality of Polar Coach."*

*"Video calls becoming an option and more widely used and accepted offering scope."*

*"Initially free access to Beam platform. Online resources (exercise videos and apps) to refer patients to."*

*"Group video calls have meant more patients can be seen at one time, less travel for staff and peer support."*

**What questions have your patients been asking you in relation to exercise and COVID-19?**

*"Patients have accessed for support/ recommendations during telephone or video consultations for general exercise advice. Been able to supply resources for home exercise and self-management exercise strategies to maintain Health and Well Being."*

*"Is it safe to go to the gym (when open)? Can they go back to team sports?"*

*"If it is safe to go out, go to gym."*

*"Ways to exercise at home or when their usual groups and classes are cancelled. A lot of our patients rely on PE and after school clubs for exercise, so we were offering advice on other things they could do at home."*

*"Is it safe enough to run outside during the pandemic? Can I train with a mask on? Is it safe enough for me to attend a gym or leisure centre?"*

*"Nuffield health availability."*

*"Asking for ideas to exercise during lock down especially for very young children/babies."*

*"Mainly parents asking for ideas for exercise/activity when they were initially shielding in the first lockdown. What they could do at home - especially in the younger children. Ideas to keep them active. If they were allowed to the park."*

*"More uptake if online exercise and treadmills/bikes/walking and outdoor cycling."*

*"Ways to keep active whilst shielding or staying indoors."*

*"Mostly questions around home exercise due to shielding."*

*"Patients have asked for resources that they are able to use at home when they have been unable to access their preferred methods of exercise. Also, how to maintain physical activity during shielding."*

*"If they can break shielding rules to continue with their exercise programmes outside/in group activities."*

*"Are they safe to go to the park or outside for a run."*

*"Selection of home exercise equipment."*

*"Infection control in gyms, swimming baths etc."*

*"Is it safe to go outside and exercise?"*

*"Safety of gyms, exercising safely outdoors, online resource advice."*

*"Lockdown terms regarding outside exercising."*

*"Asking for ideas for resources, safety of exercising outside, going back to the gym."*

*"How safe they are to exercise outside. What alternatives they can do indoors or instead of gyms etc."*

*"What equipment to purchase for home?"*

---

**Do you have any questions with regards to exercise and COVID-19 for cystic fibrosis that you would like answering/addressing?**

---

*"When is it safe to get patients back in?"*

*"I think we have only had one small child with COVID, so limited need for more in-depth questions about how they manage after."*

*"For any patients who have has CF and COVID, what was their recovery time? How did it impact on their function? Does anyone have experience regarding rehab with CF post COVID?"*

*"What types of exercise testing other centres did on patients who were at home/done remotely."*

| **Do you have any final comments on exercise and COVID-19 in your centre?** |
|---|
| *"Been fortunate that the inability to complete routine exercise testing has not hindered present clinical decision making for our cohort. However, I am uncertain as to the longer term impact of stakeholder engagement regarding commencing CPET for our cohort."* |
| *"COVID-19 has been a mixed bag for our kids - more time at home means less exposure to pathogens which they seem to have benefited from. My main concern is that this has led to a great reduction in general activity levels, combined with shielding meaning they'll be much less fit."* |
| *"Increase in numbers of patients who were exercising with us during the pandemic, i.e. better engagement than pre-pandemic perhaps as gyms closed or because of having more time. Potentially some of the changes that have happened due to the pandemic would have happened anyway but COVID has sped this up/eased the "red tape" around group virtual exercise in CF and pushed aside barriers we had."* |
| *"We share our paediatric gym with other paediatric specialities and therefore have had to come up with a timetable for use. Due to COVID-19/infection control we now have set times when we use the gym for our CF inpatients. This is continuing for the foreseeable future."* |
| *"Feel we have under provided in relation to other centres and feel this lies with the lack of confidence in building these exercise programmes -knowing where to start."* |
| *"When will we be able to confidently resume CPET?"* |
| *"If patients that are inpatients are being COVID-swabbed, why can't they use the gym as prior to pandemic if definitely negative?"* |
| *"It's really enjoyable, use of Apps for personal data collection and for exercise has increased because of COVID."* |
| *"Any guidelines for PPE required for exercise testing/guidelines of when we should resume routine exercise testing."* |
| *"We have been keen to do more but have been limited by space and time allocated to deliver this service with re-deployed staff etc."* |
| *"We plan to restart home visits for annual review ASAP in order to carry out exercise testing. With new modulator drugs prescribed for many of our patients we are unaware what has happened to exercise abilities with patients not engaging with video sessions."* |

Responses are provided as written by respondents, with only spelling mistakes and abbreviations changed to increase readability. A number of single word responses (e.g. *"stopped"*, *"videos"*) have been omitted within the Supplementary File, but counted in statistics provided in main manuscript. Free-text responses with identifying information have been removed.
